# Sensitivity of different RT-qPCR solutions for SARS-CoV-2 detection

**DOI:** 10.1101/2020.06.23.20137455

**Authors:** Julia Alcoba-Florez, Helena Gil-Campesino, Diego García-Martínez de Artola, Rafaela González-Montelongo, Agustín Valenzuela-Fernández, Laura Ciuffreda, Carlos Flores

**Author notes:** **Correspondence:** Carlos Flores, PhD., Unidad de Investigación, Hospital Universitario N.S. de Candelaria, Carretera del Rosario s/n, 38010 Santa Cruz de Tenerife.

## Abstract

**Objective:** The ongoing COVID-19 pandemic continues imposing a demand for diagnostic screening. In anticipation that the recurrence of outbreaks and the measures for lifting the lockdown worldwide may cause supply chain issues over the coming months, we assessed the sensitivity of a number of one-step retrotranscription and quantitative PCR (RT-qPCR) solutions to detect SARS-CoV-2.

**Methods:** We evaluated six different RT-qPCR alternatives for SARS-CoV-2/COVID-19 diagnosis based on standard RNA extractions. That of best sensitivity was also assessed with direct nasopharyngeal swab viral transmission medium (VTM) heating, overcoming the RNA extraction step.

**Results:** We found a wide variability in the sensitivity of RT-qPCR solutions that associated with a range of false negatives from as low as 2% (0.3-7.9%) to as much as 39.8% (30.2-50.2). Direct preheating of VTM combined with the best solution provided a sensitivity of 72.5% (62.5-81.0), in the range of some of the solutions based on standard RNA extractions.

**Conclusions:** We evidenced sensitivity limitations of currently used RT-qPCR solutions. Our results will help to calibrate the impact of false negative diagnoses of COVID-19, and to detect and control new SARS-CoV-2 outbreaks and community transmissions.

## Introduction

The ongoing pandemic of Severe Acute Respiratory Syndrome Coronavirus 2 (SARS-CoV-2) causing the coronavirus disease 2019 (COVID-19) has imposed an increasing demand for daily diagnostic screening. This is expected to perpetuate over the coming months due to the recurrence of outbreaks and the lifting of lockdown measures worldwide (Patel *et al*. 2020). Given the high sensitivity compared to serological testing (Cassaniti *et al*. 2020), standard diagnosis continues to rely on RNA extractions from respiratory or oral samples followed by one-step reverse transcription and real-time quantitative PCR (RT-qPCR) that entail one or several primer-probe sets for targeting SARS-CoV-2 sequences (Corman *et al*. 2020). While it has been shown that protocol modifications aiming to overcome supply chain issues and accelerate diagnosis affect assay sensitivity (Alcoba-Florez *et al*. 2020; Esbin *et al*. 2020), differences in target priming efficiencies and RT-qPCR kit components are also expected to account for dissimilarities in false negative results (Nalla *et al*. 2020).

Here we aimed to evaluate the sensitivity of six different RT-qPCR solutions, including five marketed kits and one based on the World Health Organization diagnostic assays with the best sensitivity (Corman *et al*. 2020; Vogels *et al*. 2020), using RNA extractions from nasopharyngeal swab viral transmission medium (VTM). The alternative with the best sensitivity was also assessed by a direct preheating of VTM samples to skip the RNA extraction step that was described elsewhere (Alcoba-Florez *et al*. 2020).

## Materials and Methods

The study was conducted at the University Hospital Nuestra Señora de Candelaria (Santa Cruz de Tenerife, Spain) from March to June 2020. We evaluated six different RT-qPCR solutions (**Table 1**), four based on three viral targets and two based on one viral target. Given the high specificity of the RT-qPCR (Alcoba-Florez *et al*. 2020), we focused on evaluating the rate of false negatives (FN) and assay sensitivity using the same 98 COVID-19 patient samples. The alternative with the best sensitivity was also assessed under an alternative procedure that skips the RNA extraction step described elsewhere (Alcoba-Florez *et al*. 2020).

**Table 1.** Different RT-qPCR solutions evaluated for the detection of SARS-CoV-2 in nasopharyngeal swab samples from COVID-19 positive patients.

| Solution | #Targets<sup>a</sup> | Target gene | Sensitivity, % (95%CI)<sup>b</sup> | FN<sup>c</sup> |
| --- | --- | --- | --- | --- |
| TaqMan Fast Virus 1-Step Master Mix (Thermo Fisher Scientific) combined with validated primer-probe sets<sup>d</sup> | 3 | E | 95.9 (89.9-98.9) | 4 |
|  |  | N | 75.5 (65.8-83.6) | 24 |
|  |  | RdRp | 77.6 (68.0-85.4) | 22 |
| LightMix® Modular SARS-CoV (COVID19) (TIB MOLBIOL) | 3 | E | 97.9 (92.8-99.7) | 2 |
|  |  | N | 78.6 (69.1-86.2) | 21 |
|  |  | RdRp | 89.8 (82.0-95.0) | 10 |
| SARS-COV-2 R-GENE (Biomérieux) | 3 | E | 65.3 (55.0-74.6) | 34 |
|  |  | N | 66.3 (56.1-75.6) | 33 |
|  |  | RdRp | 60.2 (49.8-70.0) | 39 |
| TaqPath COVID-19 CE-IVD RT-PCR Kit (Thermo Fisher Scientific) | 3 | ORF1ab | 65.3 (55.0-74.6) | 34 |
|  |  | S | 70.4 (60.3-79.2) | 29 |
|  |  | N | 76.5 (66.9-84.5) | 23 |
| Genesig Real-Time PCR COVID-19 kit (Primedesign Ltd.) | 1 | RdRp | 81.6 (72.5-88.7) | 18 |
| Real Accurate Quadruplex corona-plus PCR Kit (PathoFinder) | 1 | N | 83.7 (74.8-90.4) | 16 |

<sup>a</sup> Specific primer-probes for SARS-CoV-2.
<sup>b</sup> 95% Confidence Interval.
<sup>c</sup> False negative counts out of 98 patients.
<sup>d</sup> Corman *et al.* (2020).

Samples were collected in 2 mL of VTM (Biomérieux). RNA extractions were conducted from 200 μL of VTM using the MagNA Pure Compact Nucleic Acid Isolation Kit I (Roche) or the STARMag Viral DNA/RNA 200C kit (Seegene). The RT-qPCR was performed in 10 μL final volume reactions (5 μL of sample) using a CFX96 Touch Real-Time PCR Detection System (Bio-Rad) following the thermal cycling specifications of each solution. Positive and negative controls were included in all experiments as described elsewhere (Alcoba-Florez *et al*. 2020). Sensitivity and 95% confidence intervals (95% CI) were calculated from the FN counts using MedCalc (MedCalc Software Ltd.).

## Results

Since all samples were COVID-19 positive for at least one solution/viral target, results with threshold cycle (Ct) values above 40 or those that remained undetected during the 45 cycles of the experiments were considered FN observations (**Figure 1, Table 1**).

**Figure 1.**
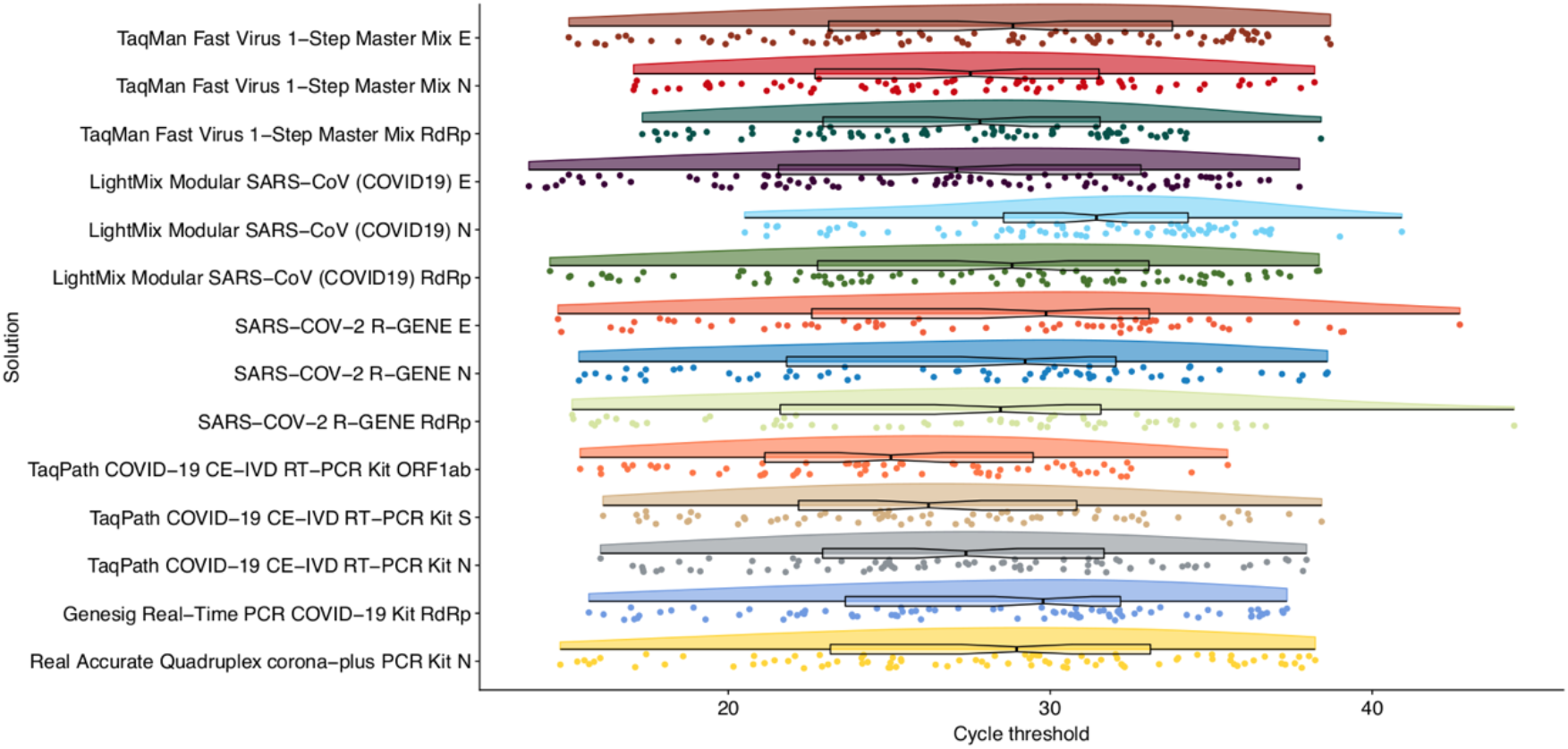
Raincloud plot of the distribution of cycle threshold (Ct) values for the RT-qPCR solutions evaluated for the detection of SARS-CoV-2 in COVID-19 positive samples. Raw Ct data with the median and the interquartile range are also represented overlaid on each distribution.

Attending to individual targets, we found that the most sensitive solution was the LightMix® Modular SARS-CoV (COVID19) (TIB MOLBIOL) used in combination with a primer-probe set for the E-gene (97.9% [92.8-99.7]) (**Table 1**). It was followed closely by the TaqMan Fast Virus 1-Step Master Mix (Thermo Fisher Scientific) kit combined with validated primer-probes for diagnosis (Corman *et al*. 2020) for the same viral gene (95.9% [89.9-98.9]). When combining at least two viral gene targets, we found that the TaqMan Fast Virus 1-Step Master Mix kit with validated primer-probe sets targeting both E and RdRp genes (Corman *et al*. 2020) attained an equivalent sensitivity. The kit with the poorest performance for all the three viral primer-probe sets was SARS-COV-2 R-GENE (Biomérieux) (range of 60.2% [49.8-70.0] to 66.3% [56.1-75.6]). Its levels of sensitivity improved to those of all other kits when the E-gene primer-probe set was combined with those for N or the RdRp genes (71.4% [61.4-80.1] and 69.4% [59.3-78.3], respectively). The sensitivity of all other solutions did not benefit from combining the result of more than one primer-probe set.

Finally, because the LightMix® Modular SARS-CoV (COVID19) kit with primer-probes for the E-gene showed the highest sensitivity, we tested it on samples that were preheated at 70°C for 10 min in substitution of the RNA extraction (Alcoba-Florez *et al*. 2020). Although this alternative decreased the kit sensitivity (72.5% [62.5-81.0]), the results were still comparable to other evaluated solutions (**Table 1**).

## Discussion

RT-qPCR for selected target genes of SARS-CoV-2 has been key in the global response to the pandemic. Given the rapid spread of the virus at this time, it is likely that the RT-qPCR assays will continue to be a central tool for controlling COVID-19. However, as happened in the past due to supply chain issues, policy decisions and laboratory testing capacities (Alcoba-Florez *et al*. 2020), it is predictable that the diagnosis of COVID-19 will continue relying on a variety of solutions among laboratories and countries (Vogels *et al*. 2020).

Our results evidenced a wide variability in the sensitivity of RT-qPCR solutions for SARS-CoV-2 detection which associated with a proportion of FN ranging from as low as 2% (0.3-7.9%) to as much as 39.8% (30.2-50.2). These findings will help to assess the impact of the selected solution on FN diagnoses of COVID-19 (Ramdas *et al*. 2020) and to choose a solution that minimize misdiagnoses of an active SARS-CoV-2 infection.

## Data Availability

Not applicable.

## Authors’ contributions

JAF and CF designed the study. JAF, HGC, and DGM participated in data acquisition. JAF, LC and CF performed the analyses and data interpretation. LC, AVF, RGM and CF wrote the draft of the manuscript. All authors contributed in the critical revision and final approval of the manuscript.

## Acknowledgments

We deeply acknowledge the University Hospital Nuestra Señora de Candelaria board of directors and the executive team for their strong support and assistance in accessing diverse resources used in the study.

## Conflicts of Interest

The authors declare that they have no known competing financial interests or personal relationships that could have appeared to influence the work reported in this paper.

## Funding

This research was funded by Cabildo Insular de Tenerife [grant number CGIEU0000219140]; the agreement with Instituto Tecnológico y de Energías Renovables (ITER) to strengthen scientific and technological education, training research, development and innovation in Genomics, Personalized Medicine and Biotechnology [grant number OA17/008]; Ministerio de Innovación y Ciencia [grant number RTI2018-093747-B-100 and RTC-2017-6471-1], co-funded by the European Regional Development Fund (ERDF); Lab P2+ facility [grant number UNLL10-3E-783], co-funded by the ERDF and “Fundación CajaCanarias”; and the Spanish HIV/AIDS Research Network [grant number RIS-RETIC, RD16/0025/0011], co-funded by Instituto de Salud Carlos III and by the ERDF.

## Ethical Approval

The University Hospital Nuestra Señora de Candelaria (Santa Cruz de Tenerife, Spain) review board approved the study (ethics approval number: CHUNSC_2020_24).

## Notes

### Competing Interest Statement

The authors have declared no competing interest.

